# Novel molecular biomarkers to guide treatment-decision making in metastatic urothelial cancer - A patient cohort analysis

**DOI:** 10.1101/2025.04.02.25325155

**Authors:** Debbie G.J. Robbrecht, Youssra Salhi, John W.M. Martens, Maureen J. B. Aarts, Paul Hamberg, Michiel S. van der Heijden, Jens Voortman, Niven Mehra, Hans M. Westgeest, Ronald de Wit, Reno Debets, Joost L. Boormans, J. Alberto Nakauma-González

## Abstract

The current options and recent developments in the field of systemic therapy for advanced urothelial cancer (UC) patients, urges the need for selection criteria to identify the most optimal therapeutic option for individual patients. The molecular makeup of tumors, including molecular subtype, tumor microenvironment composition, gene mutations, fusions and amplifications, has been previously correlated with response to immune checkpoint inhibitors (ICI), erdafitinib or EV monotherapy respectively, and may withhold potential candidate biomarkers. In this study, we aimed to stratify mUC patients based on molecular biomarkers that might be associated with response to EV, an FGFR inhibitor, or anti-PD-(L)1, by using whole-genome DNA- and paired RNA-sequencing data of fresh-frozen metastatic tumor biopsies of 155 mUC patients. We observed that *NECTIN4* amplification, *FGFR2/3* mutations, and the RNA-expression-based T-Cell-to-Stroma Enrichment (TSE) score were mutually exclusive, and may therefore reflect biologically distinct tumors and sensitivity to treatments. This finding was validated in two independent bladder cohorts: the IMvigor210 study and The Cancer Genome Atlas. Stratification of patients into subgroups based on these molecular features is possible.

Our data challenge the concept of a one-treatment-fits-all paradigm and support the rationale for prospective clinical trials with biomarker-guided treatment selection of mUC patients.

## Main text

The treatment landscape of locally advanced or metastatic urothelial carcinoma (mUC) has changed since the introduction of immune checkpoint inhibitors (ICI), antibody-drug-conjugates (ADCs), and targeted therapies. In recent years, the U.S. Food and Drug Administration (FDA) approved the ADC enfortumab vedotin (EV) as a monotherapy in patients who progressed after prior treatment with platinum-based chemotherapy and PD-(L)1 ICI, or in case cisplatin-ineligibility, after prior PD-(L)1 ICI; and more recently, the combination of EV with pembrolizumab (EVP) in the first-line setting. Also the FGFR inhibitor erdafitinib has been approved for mUC patients whose tumors harbor susceptible FGFR3 gene alterations and who progressed after 1 or 2 prior therapies, including an PD-(L)1 ICI. With more therapeutic options available, there is an urgent need for predictive biomarkers to improve patient stratification, optimize treatment of choice in 1^st^ and subsequent lines, and omit ineffective therapies in patients who are unlikely to benefit. Also, given the large burden of novel therapies on the healthcare system and financial toxicity, efficient decision-making is key.

Predictive biomarkers to guide treatment-decision making are *FGFR2/3* mutations or fusions for the treatment with FGFR inhibitors [1] and PDL1 expression for anti-PD(L)1 ICI in first-line setting in cisplatin ineligible patients [2, 3]. The response to ICI depends on, i.a., the tumor microenvironment. We recently demonstrated that the T cell-To-Stroma enrichment (TSE) score, a metric that estimates the relative abundance of T cells over stromal cells in tumor samples, corresponded to response to ICI [4]. A positive TSE score at baseline favored response to pembrolizumab and these patients lived twice as long as patients without a positive TSE in their tumor samples. For EV treatment, there is no selection criterion and all patients sufficiently fit could be treated per its label. A recent study, however, showed that 96% of mUC patients with *NECTIN4* amplification (~25% of all patients) showed objective responses to EV compared with 32% in the nonamplified subgroup, indicating a potential role for *NECTIN4* amplification in treatment decision making [5]. In view of the mechanism of EV, it is appealing to consider a role of the tumor microenvironment and therefore the TSE score, in correlation with EV efficacy. Though this is yet unknown.

In this study, we analyzed *NECTIN4* amplification, the TSE score and *FGFR2/3* mutations in a cohort of 155 locally advanced and mUC patients who were scheduled for palliative systemic treatment (discovery cohort). In 74 patients, tumor samples were collected prior to first or second-line ICI monotherapy, and 87 patients received chemotherapy, combination chemo-immunotherapy or other therapies (see Table S1 for patient characteristics). No patients received erdafitinib, EV or EVP.

Using whole-genome DNA- and paired RNA-sequencing data of fresh-frozen metastatic tumor biopsies of 155 mUC patients [4,6] we found that *NECTIN4* amplification, *FGFR2/3* mutations and fusions, and the positive TSE score were mutually exclusive (Figure 1A-B; p ≤ 0.001 in all cases, estimations using the Poisson-Binomial distribution). This finding might be explained because both *NECTIN4* and *FGFR3* are drivers of UC [6], and may represent different tumor biology. Tumors with a positive TSE score showed the highest expression of the immune checkpoint genes *PDCD1, CD274* and *CTLA4* compared to tumors with *NECTIN4* amplification, *FGFR2/3* mutations and tumors lacking these mutations (Figure 1C; q < 0.001 in all cases, Kruskal-Wallis test). Tumors with *NECTIN4* amplification were enriched for the luminal molecular subtype and had the highest RNA expression of *NECTIN4* (q < 0.001, Kruskal-Wallis test). Amplifications in *TACSTD2* (TROP-2), another potential target for therapy [7], were nearly absent but those few cases present overlapped with cases having *NECTIN4* amplification.

**Figure 1.**
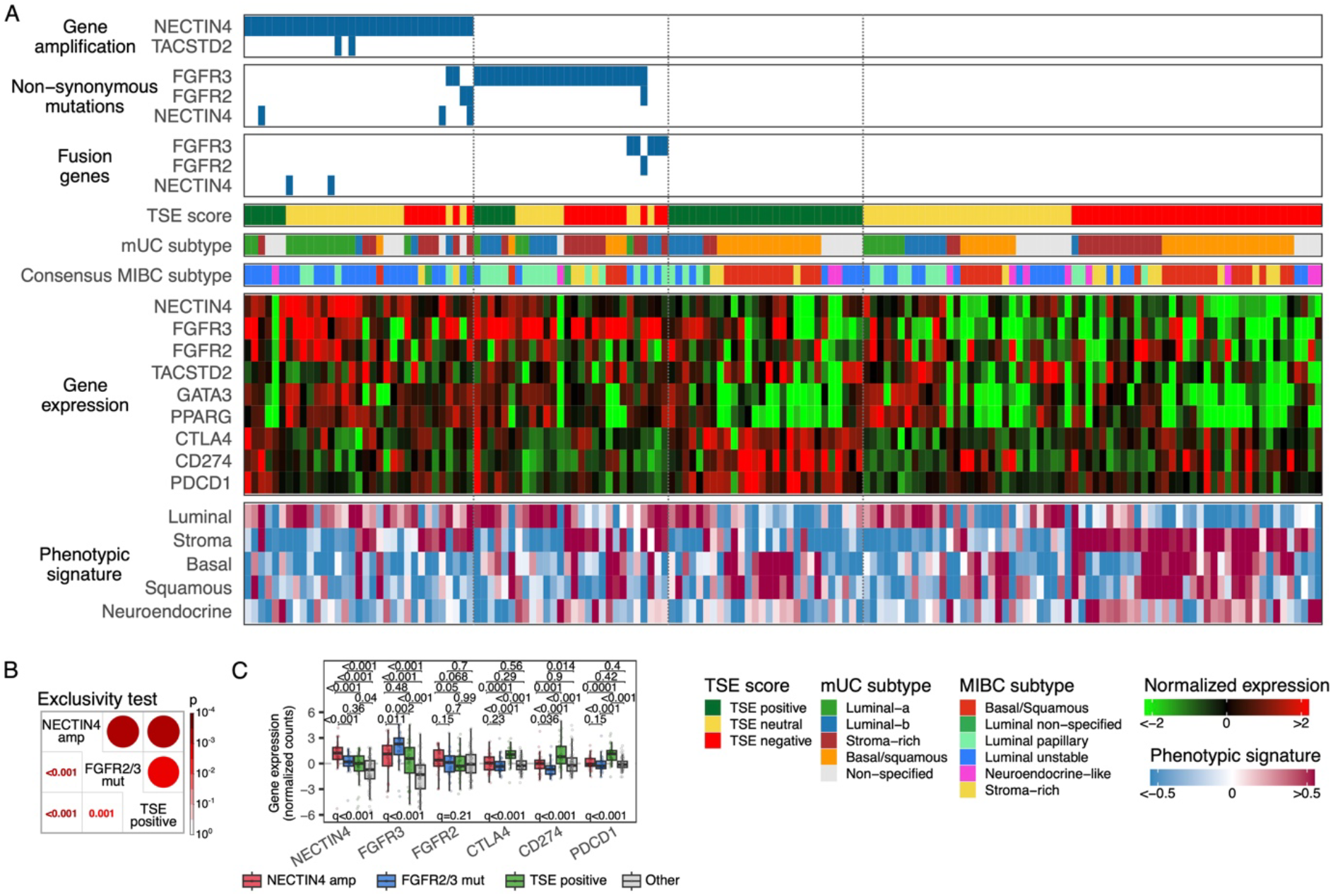
Molecular alterations in metastatic urothelial carcinomas (mUC) that might serve as predictive biomarkers to systemic therapy. **A)** From top to bottom: gene amplifications; non-synonymous single nucleotide variants; fusion genes; the RNA-based T cell-To-Stroma enrichment (TSE) score; mUC molecular subtype; consensus muscle-invasive bladder cancer (MIBC) molecular subtype; RNA expression of selected genes; and phenotypic signatures. **B)** Exclusivity test between *NECTIN4* amplifications, *FGFR2/3* genomic alterations and the positive TSE score. **C)** Expression of selected genes across tumors with *NECTIN4* amplification, *FGFR2/3* mutations (including fusions) and positive TSE score. A pair-wise Wilcoxon rank-sum test was applied on the gene expression between groups and p-values were not corrected (top of boxplots). Additionally, the Kruskal-Wallis test was applied and q values are the adjusted p values using the Benjamini-Hochberg corrected method (bottom of boxplots).

Mutual exclusivity between *FGFR2/3* mutations and a positive TSE score was validated in the independent cohort of the IMvigor210 study (Figure S1; p < 0.001 estimated using the Poisson-Binomial distribution). Amplification of *NECTIN4* was not available in the dataset of this cohort, however, *NECTIN4* amplification estimated from RNA expression levels confirmed the mutually exclusive pattern between the three biomarkers (Figure S2). In the Cancer Genome Atlas (TCGA) bladder cancer cohort, mutual exclusivity of *NECTIN4* amplification, *FGFR2/3* mutations, and a positive TSE score was also confirmed for primary non-metastatic muscle-invasive urothelial carcinoma of the bladder (Figure S3; p < 0.001 in all cases, estimations using the Poisson-Binomial distribution).

Based on these data, stratification into four subgroups is possible; a) Patients with *NECTIN4* amplified tumors enriched for the luminal subtype. These patients might have most benefit from EV as monotherapy, without the need to combine with pembrolizumab, b) patients with *FGFR2/3* mutations/fusions and no *NECTIN4* amplification might have most benefit from primary treatment with an FGFR inhibitor, c) patients with a positive TSE score without *NECTIN4* amplification or *FGFR2/3* mutations/fusions might benefit most from anti-PD(L)1 ICI monotherapy, and d) in patients with a neutral or negative TSE score, lack of *NECTIN4* amplification, and no *FGFR 2/3* mutations/fusions but enriched for the basal and stroma-rich molecular subtype, the most optimal therapeutic strategy is unclear based on these biomarkers (Figure 2).

**Figure 2.**
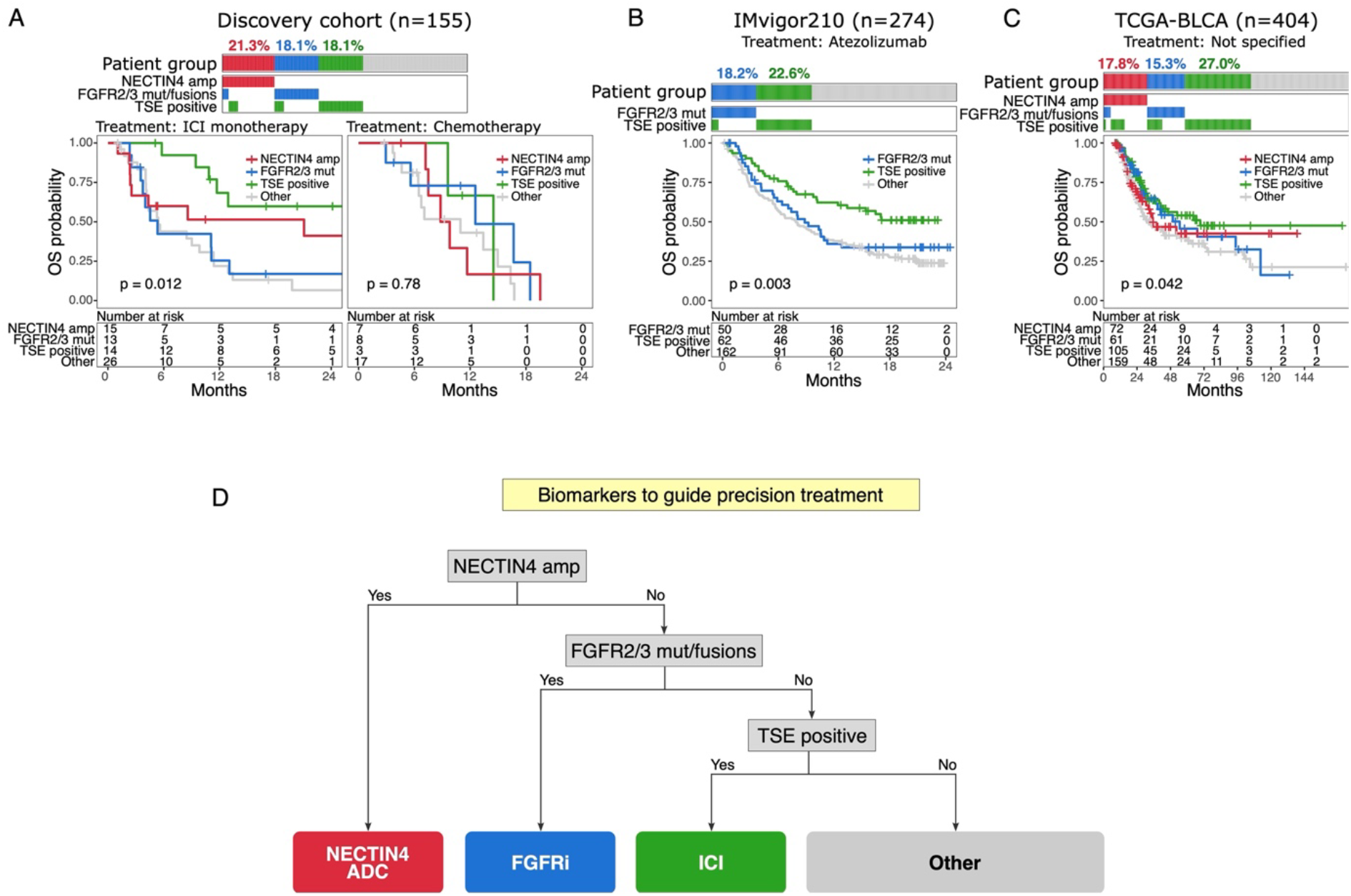
Biomarkers to guide targeted therapy in metastatic urothelial carcinoma. Overall survival of patients with metastatic (panel A Discovery cohort; and B IMvigor210 cohort) and primary (panel C TCGA cohort) urothelial carcinoma. Patient populations were stratified based on *NECTIN4* amplification, *FGFR2/3* mutations/fusions (no *NECTIN4* amplification) and TSE score (no *FGFR2/3* mutations and no *NECTIN4* amplification): panel **A)** shows survival outcomes of mUC patients with available follow up from the Discovery cohort who were treated with ICI monotherapy (N=68) or chemotherapy (N=35), panel **B)** shows survival outcomes of mUC patients from the IMvigor210 study who were treated with atezolizumab, and panel **C)** shows the survival outcomes of patients with primary urothelial carcinoma of the bladder from the TCGA cohort. **D)** Flowchart for biomarker-guided clinical trials in patients with mUC. mUC = metastatic urothelial carcinoma; amp = amplification, mut = mutation/fusion

Although the THOR trial showed no superiority of erdafitinib in second line [8], compared to pembrolizumab in patients with PDL1 naïve tumors, it does not reject the hypothesis of the presence of a subgroup of patients with best anticipated response from upfront FGFR inhibition, based on a combination of molecular features of their tumors.

A recent study found that most drug combination therapies (95%) approved for advanced cancers exhibited additive or less than additive effects [9]. This suggests that only in a minority of patients where we observed overlapping biomarkers may benefit from combined therapies and the added value of drug combinations in other cases may be negligible. Further studies are needed to investigate whether EV and pembrolizumab have a synergistic effect or both drugs target different subpopulations within one population.

Based on the biomarkers analyzed in this study, we propose a biomarker-guided stratification flowchart (Figure 2D), that could serve as a rationale for biomarker-guided clinical trials. Subgroups are defined by the presence or absence of 1) *NECTIN4* amplification, 2) *FGFR2/3* alteration, 3) positive TSE score. Patients from the cohorts who might have derived most benefit from PD(L)1 ICI by using the biomarker-guided stratification flowchart (TSE positive score, no *NECTIN4* amplification, no *FGFR2/3* alteration) and who actually received ICI monotherapy (discovery and Imvigor210 cohort), indeed had a longer overall survival (Figure 2A-B and Figure S2C), and was independent of primary tumor location, metastatic biopsy site, age and sex (Figure S4).

Interestingly, clinical outcomes in patients were comparable between the different biomarker-guided stratified subgroups when treated with platinum-based chemotherapy (Figure 2A) or in case of primary non-metastatic bladder cancer (no treatment with systemic therapy) in the TCGA-BLCA cohort (Figure 2C).

It is important to note that this study has an exploratory nature and that several limitations might have impacted the outcomes of the analyses; Data from the discovery cohort are based on a relatively small sample size and both patient population (primary tumor location and biopsy site) and applied (previous) treatments are heterogeneous. Another limitation is the lack of patients treated with EV or combination therapies, like EV plus pembrolizumab. Therefore, more prospective studies are needed to address biomarker development for treatment, especially in first-line setting. Despite the limitations of our study, it gives an important awareness signal and may feed future initiatives in optimizing use of available resources next to optimizing care and outcomes of patients.

In conclusion, our exploratory study on *NECTIN4* amplification, *FGFR2/3* genomic alterations and the RNA-based TSE score shows the potential of biomarker-guided therapy for personalized medicine in advanced urothelial carcinoma patients, challenges the concept of a one-size-fits all treatment strategy and supports the rationale for prospective clinical trials with biomarker-guided treatment selection of mUC patients.

## Data Availability

All data are freely available for academic use from the Hartwig Medical Foundation at https://www.hartwigmedicalfoundation.nl/en/data/data-access-request/.

https://portal.gdc.cancer.gov

http://research-pub.gene.com/IMvigor210CoreBiologies/

## Acknowledgements

The Hartwig Medical Foundation and the Center of Personalized Cancer Treatment are acknowledged for making the clinical and genomic/transcriptomic data available to the study. We thank all local principal investigators and the nurses of all contributing centers for their help with patient recruitment. We are particularly grateful to all participating patients and their families.

