## Supplementary for "Novel molecular biomarkers to guide treatment-decision making in metastatic urothelial cancer - A patient cohort analysis"

Debbie G.J. Robbrecht, *et al*.

Department of Medical Oncology, Erasmus MC Cancer Institute, University Medical Center Rotterdam, Rotterdam 3015 GD, the Netherlands

### Methods

#### Data and code availability

All code and scripts are available at https://github.com/hartwigmedical/ and at https://github.com/ANakauma/biomarkers-guided-treatment-mUC/.

All pre- and post-processed WGS data, RNA-seq data and corresponding clinical data have been requested from the Hartwig Medical Foundation (HMF) and were provided under data request number DR-314. All data are freely available for academic use from the HMF through standardized procedures. Request forms can be found at https://www.hartwigmedicalfoundation.nl.

#### Patient cohort

Patients with advanced or metastatic urothelial carcinoma (mUC) were prospectively enrolled in several multicenter clinical trials coordinated with the Hartwig Medical Foundation (HMF). These patients were scheduled for 1st or 2nd line palliative systemic treatment. Overall survival was defined as the time from start of treatment until death. Survivors were censored at the last date of response evaluation. Following protocols of HMF [1], whole-genome DNA-sequencing (WGS) with a depth close to 100X, was successfully performed on 155 samples from freshly obtained biopsies from metastatic sites, and matched RNA-sequencing (RNA-seq) was available for all these samples.

**WGS and RNA-seq data analysis**

Alignment and pre-processing of WGS data was performed by the HMF using the hg19 reference genome [1]. Structural variants, including copy number and fusion genes, estimated from WGS by algorithms developed by the HMF (https://github.com/hartwigmedical/hmftools) were also provided. Copy numbers were used to estimate significant deep amplifications of genes with GISTIC2 v2.0.23.

Alignment and pre-processing of RNA-seq data using the hg19 reference genome have been previously described [2]. RNA counts were estimated using Kallisto v0.48.0. The transcriptomic subtype of each mUC sample was identified using the mUC classifier (https://bitbucket.org/ccbc/dr31_hmf_muc). The TSE score was estimated by applying the TSE classifier (https://github.com/ANakauma/TSEscore_ICIs). A list of genes associated to each phenotype [2] was used to calculate the signature score with the Gene Set Variation Analysis method (GSVA v1.48.0).

**Statistical analysis**

Analyses were performed using the statistical analysis platform R v4.3.2 [3]. The Kruskal-Wallis test was used for comparison between groups and p values were adjusted using the Benjamini-Hochberg method. Pair-wise comparisons were performed with the Wilcoxon rank-sum test. The Poisson-binomial method was applied for mutually exclusive mutation events using Rediscover v0.3.2 [4]. The log-rank test was used for comparing Kaplan–Meier survival curves and the multivariate Cox proportional hazards regression analysis was applied to overall survival.

### Supplementary Tables/Figures

Table S1. Characteristics of 155 patients with metastatic urothelial cancer stratified by treatment with immune checkpoint inhibition versus chemotherapy.

| Discovery cohort characteristics | Total cohort | Patient with follow up after treatment | |
|---|---|---|---|
| | | Patients treated with ICI | Patients treated with chemotherapy |
| | N = 155 | N = 68 | N = 35 |
| Age, years | | | |
| Median (Q1-Q3) | 68 (61-73) | 70 (64-73) | 66 (61-71) |
| Unknown | 12 (8%) | | |
| Sex | | | |
| Female | 34 (22%) | 15 (22%) | 8 (23%) |
| Male | 110 (71%) | 53 (78%) | 27 (77%) |
| Unknown | 11 (7%) | 0 | 0 |
| Primary tumor location | | | |
| Bladder | 115 (74%) | 57 (84%) | 22 (63%) |
| Upper tract | 21 (14%) | 7 (10%) | 7 (20%) |
| Urethra | 13 (8%) | 3 (4%) | 5 (14%) |
| Urachus | 2 (1%) | 0 | 1 (3%) |
| Upper tract and urethra | 1 (<1%) | 1 (1%) | 0 |
| Unknown | 3 (2%) | 0 | 0 |
| Biopsy sites | | | |
| Lymph node | 51 (33%) | 24 (35%) | 16 (46%) |
| Liver | 40 (26%) | 14 (21%) | 11 (31%) |
| Bone | 8 (5%) | 3 (4%) | 1 (3%) |
| Lung | 8 (5%) | 5 (7%) | 2 (6%) |
| Other | 36 (23%) | 22 (32%) | 5 (14%) |
| Unknown | 12 (8%) | 0 | 0 |
| Systemic pretreatment | | | |
| Yes | 93 (60%) | 62 (91%) | 8 (23%) |
| No | 47 (30%) | 6 (9%) | 27 (77%) |
| Unknown | 15 (10%) | 0 | 0 |
| Radiotherapy pretreatment | | | |
| Yes | 42 (27%) | 18 (26%) | 9 (26%) |
| No | 98 (63%) | 50 (74%) | 26 (74%) |
| Unknown | 15 (10%) | 0 | |
| Treatment received after biopsy | | | |
| Pembrolizumab | 68 (44%) | 62 (91%) | 0 |
| Gemcitabine, cisplatin and/or carboplatin | 33 (21%) | 0 | 26 (74%) |
| Chemotherapy in combination with ICI | 5 (3%) | 0 | 4 (11%) |
| Nivolumab | 3 (2%) | 3 (4%) | 0 |
| Atezolizumab | 2 (1%) | 2 (3%) | 0 |
| Tremelimumab | 1 (<1%) | 1 (1%) | 0 |
| Other agents and/or combinations | 7 (5%) | 0 | 5 (14%) |
| Unknown | 36 (23%) | 0 | 0 |
| Follow-up after treatment | | | |
| Yes | 105 (68%) | 68 (100%) | 35 (100%) |
| No | 50 (32%) | | |
| Follow-up, months | | | |
| Median (Q1-Q3) | 8.4 (4.1-13.4) | 7.1 (3.9-14.3) | 8.8 (6.0-12.6) |
| Mean ± SD | 10.4 ± 8.1 | 10.7 ± 9.2 | 9.5 ± 4.7 |

| | | | |
|---|---|---|---|
| Follow up for survivors, months | | | |
| Median (Q1-Q3) | 10.9 (5.1-22.2) | 17.0 (5.2-25.3) | 6.9 (4.9-10.5) |
| Survival, months | | | |
| Median (95% CI) | 10.8 (8.4-12.9) | 9.8 (5.6-12.9) | 10.9 (7.5-16.3) |

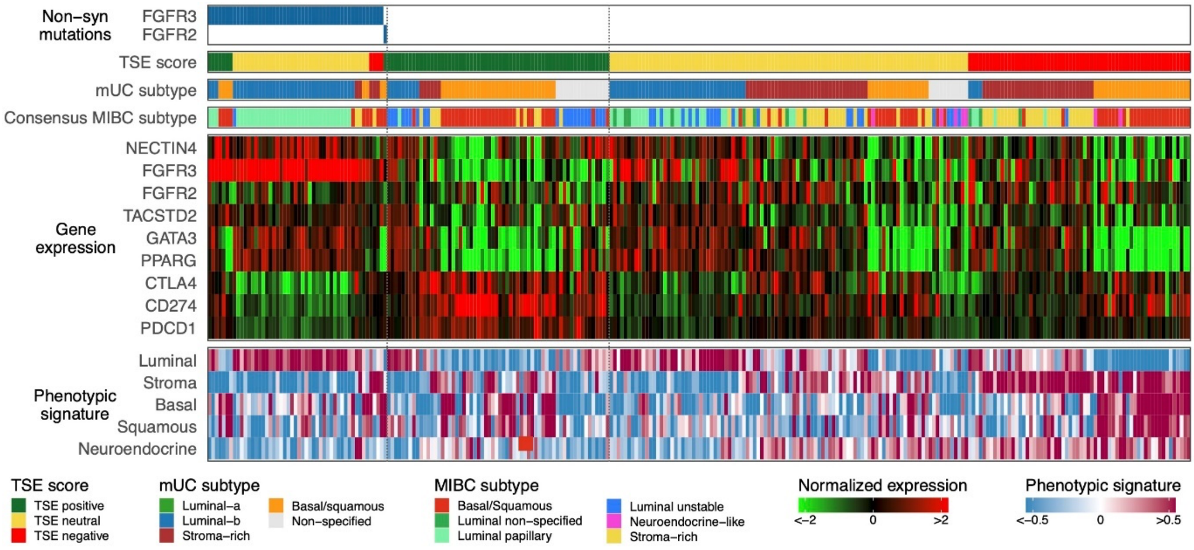


Figure S1. The molecular landscape of predictive markers to therapy in 274 metastatic urothelial carcinomas (mUC). From top to bottom: non-synonymous single nucleotide variants; the TSE score; mUC molecular subtype; consensus muscle-invasive urothelial carcinoma (MIBC) subtype; RNA expression of selected genes; and phenotypic signatures.

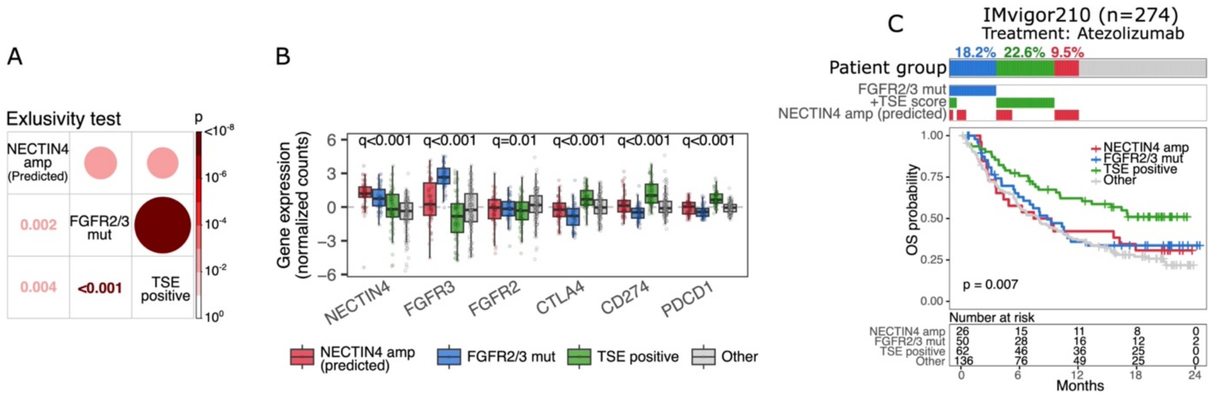


Figure S2. Predictive biomarkers in 274 metastatic urothelial carcinomas (mUC). *NECTIN4* amplification was estimated from RNA using CNVkit (https://github.com/etal/cnvkit) and the correlation between expression levels and amplification from the TCGA bladder cancer cohort. **A)** Exclusivity test between samples with predicted *NECTIN4* amplifications, *FGFR2/3* genomic alterations and the positive TSE score. **B)** Expression of selected genes across tumors with predicted *NECTIN4* amplification, *FGFR2/3* mutations and positive TSE score. The Kruskal-Wallis test was applied and q values are the adjusted p values using the Benjamini-Hochberg corrected method. **C)** Copy number estimation from RNA is usually less accurate than DNA-sequencing and for this reason samples were stratified firs by *FGFR2/3* mutations, then positive TSE score (in absence of *FGFR2/3* alterations) and finally *NECTIN4* amplification (neither *FGFR2/3* mutations or positive TSE score). The survival outcomes of these group of patients were compared with the log-rank test.

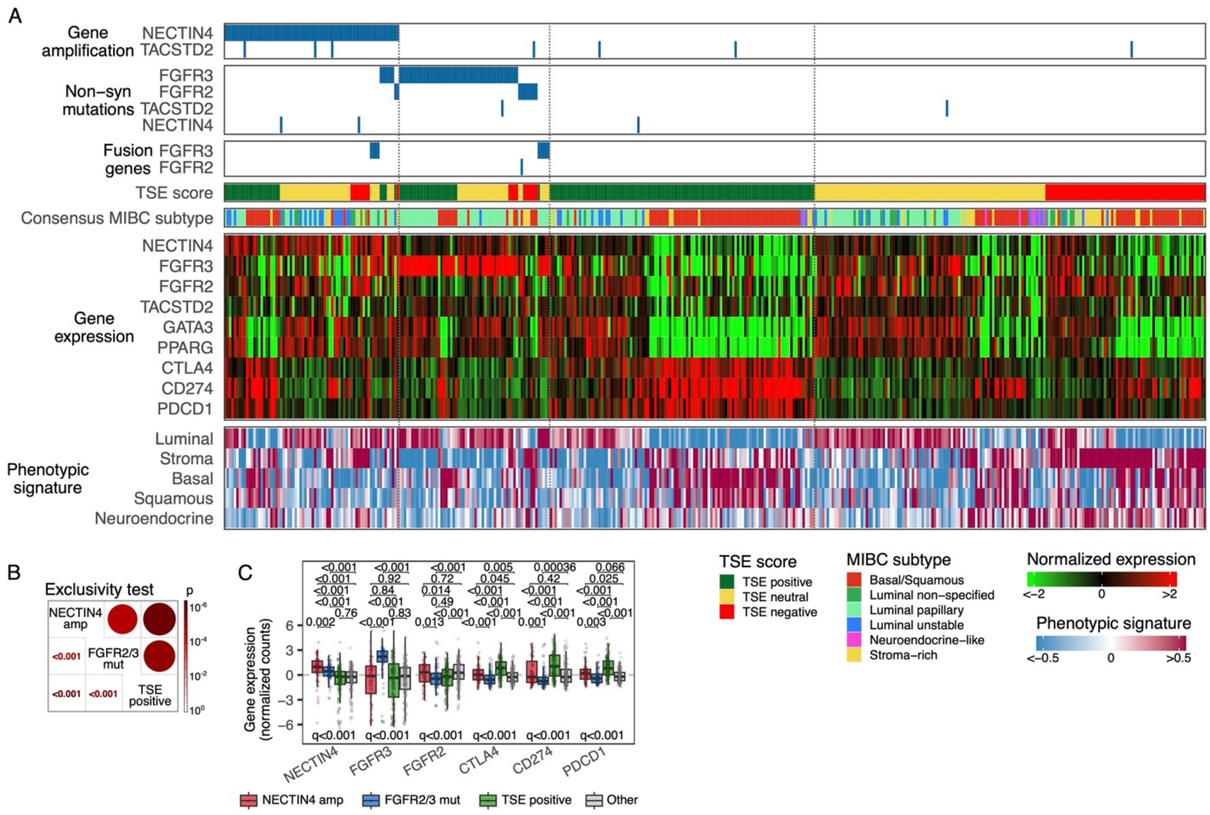


Figure S3. The molecular landscape of predictive markers to therapy in 404 primary urothelial carcinomas from the TCGA cohort. Due to the lack of other metastatic cohorts with available (complete) genomic and transcriptomic data, the TCGA dataset was used to validate the predictive biomarkers. **A)** From top to bottom: gene amplifications; non-synonymous single nucleotide variants; fusion genes; the TSE score; consensus muscle-invasive urothelial carcinoma (MIBC) subtype; RNA expression of selected genes; and phenotypic signatures. **B)** Exclusivity test between *NECTIN4* amplifications, *FGFR2/3* genomic alterations and the positive TSE score. **C)** Expression of selected genes across tumors with *NECTIN4* amplification, *FGFR2/3* mutations (including fusions) and positive TSE score. A pair-wise Wilcoxon rank-sum test was applied to the gene expression between groups and p-values were not corrected (top of boxplots). Additionally, the Kruskal-Wallis test was applied and q values are the adjusted p values using the Benjamini-Hochberg corrected method (bottom of boxplots).

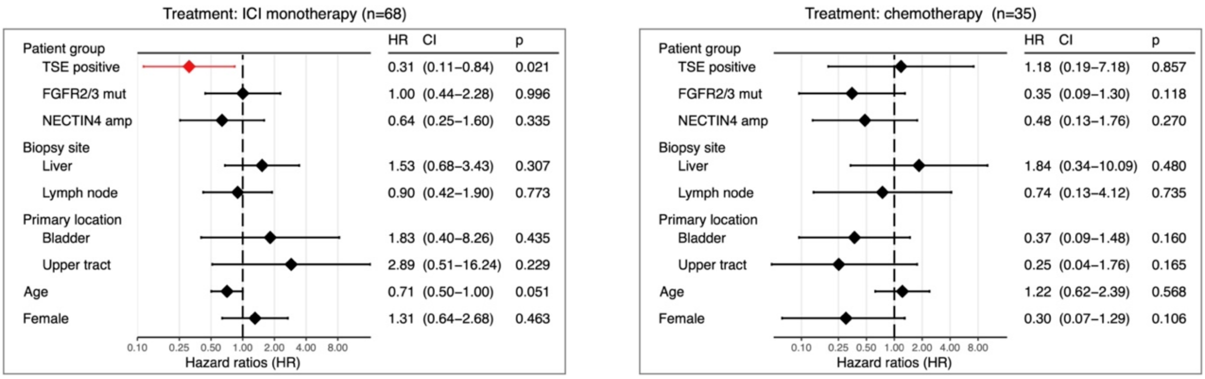


Figure S4. Multivariate Cox regression analysis for biomarker-guided stratification of patients and cohort characteristics. The Cox proportional hazards regression analysis was applied to overall survival in patients who received immune checkpoint inhibitors (ICI) or chemotherapy. All variables were categorized except for age in which rescaled (divided by 10) continuous values were used. Hazard ratios (HR) and 95% confidence intervals (CI) are displayed.
